# Genome-wide meta-analyses of non-response to antidepressants identify novel loci and potential drugs

**DOI:** 10.1101/2024.07.13.24310361

**Authors:** Elise Koch, Tuuli Jürgenson, Guðmundur Einarsson, Brittany Mitchell, Arvid Harder, Luis M. García-Marín, Kristi Krebs, Yuhao Lin, Ying Xiong, Estonian Biobank Research Team, Yi Lu, Sara Hägg, Miguel E. Rentería, Sarah E. Medland, Naomi R. Wray, Nicholas G. Martin, Christopher Hübel, Gerome Breen, Thorgeir Thorgeirsson, Hreinn Stefánsson, Kári Stefánsson, Lili Milani, Ole A. Andreassen, Kevin S. O’Connell

## Abstract

Antidepressants exhibit a considerable variation in efficacy, and increasing evidence suggests that individual genetics contribute to antidepressant treatment response. Here, we combined data on antidepressant non-response measured using rating scales for depressive symptoms, questionnaires of treatment effect, and data from electronic health records, to increase statistical power to detect genomic loci associated with non-response to antidepressants in a total sample of 135,471 individuals prescribed antidepressants. We performed genome-wide association meta-analyses, leave-one-out polygenic prediction, and bioinformatics analyses for genetically informed drug prioritization. We identified two novel loci associated with non-response to antidepressants and showed significant polygenic prediction in independent samples. In addition, we investigated drugs that target proteins likely involved in mechanisms underlying antidepressant non-response, and shortlisted drugs that warrant further replication and validation of their potential to reduce depressive symptoms in individuals who do not respond to first-line antidepressant medications. These results suggest that meta-analyses of GWAS utilizing real-world measures of treatment outcomes can increase sample sizes to improve the discovery of variants associated with non-response to antidepressants.

## Introduction

Antidepressants are the first-line pharmacological treatment for depression. Over 10% of the adolescent population uses antidepressant medication, and the rate of antidepressant prescriptions is increasing^1, 2^. Selective serotonin reuptake inhibitors (SSRIs) are the most used antidepressants^3-5^, because they are generally better tolerated compared to other antidepressant classes^4, 5^. However, treatment response to SSRIs and other antidepressants varies considerably between treated individuals, and less than half of individuals with major depression achieve remission of symptoms after initial antidepressant treatment^6, 7^. It has been shown that individuals who require several antidepressant treatment steps show worse longer-term treatment outcomes^7^. Although antidepressants are linked to a reduction in depressive symptoms^8^, they are often ineffective, with only approximately 35% achieving remission after their primary antidepressant treatment^6^, and approximately 50% achieving remission after completing two treatments of antidepressants^7^. Antidepressant non-response has been associated with illness severity, more comorbidities, higher antidepressant dose requirements, and higher suicide risk as well as suicide attempts^9, 10^. Thus, non-response to antidepressants is a major clinical problem, and early identification remains a critical priority in psychiatry research^11^.

Increasing evidence suggests that genetic variation contributes to antidepressant treatment outcomes^11^. Discovering genomic variants associated with antidepressant treatment outcomes could facilitate the early identification of individuals who do not respond to first-line treatments to avoid delay in reaching recovery and advance personal treatment. However, although common single nucleotide polymorphisms (SNPs) are reported to explain 42% of the variance of antidepressant response^12^, no robustly replicated associations have been detected to date^13-17^. Moreover, the largest genome-wide association study (GWAS) of antidepressant response, measured using depression symptom scores (N=5,218), did not identify any genome-wide significant loci^18^. Antidepressant response is a polygenic phenotype, requiring larger sample sizes to elucidate the genetic architecture of antidepressant response^18^. Use of alternative outcome phenotypes such as antidepressant response information obtained from electronic health records (eHR)^19^ or self-reported questionnaires^20^ have been used to increase sample sizes. Combining these real-world data sources could provide the sample sizes needed for discovering genetic factors associated with antidepressant treatment outcomes^21^. In the current study, we integrated GWAS data on antidepressant non-response measured using rating scales for depressive symptoms, questionnaires of treatment effect, and outcome data from eHR, to increase statistical power to detect genomic loci associated with non-response to SSRIs and serotonin-norepinephrine reuptake inhibitors (SNRIs). We identified one novel locus associated with non-response to SSRIs, and one novel locus associated with non-response to SSRIs/SNRIs, as well as a replicable polygenic signal of non-response to SSRIs. Based on bioinformatics analyses, we shortlisted approved drugs that target proteins likely involved in mechanisms underlying antidepressant non-response.

## Methods

### GWAS sample description

Using questionnaire data about the effectiveness of prescribed antidepressant drugs, we performed GWASs on non-response to SSRIs in the Estonian Biobank (EstBB)^22^, the Australian Genetics of Depression Study (AGDS)^23^, the Genetic Links to Anxiety & Depression (GLAD) Study^24^, and the UK Biobank (UKB)^25^. Additionally, we performed GWASs on non-response to SNRIs in the EstBB and AGDS cohorts. Utilizing prescription registry data, we defined treatment response and non-response to antidepressants based on antidepressant switching and performed GWASs on non-response to SSRIs and SNRIs in an Icelandic cohort from deCODE Genetics. In all cohorts, treatment response and non-response was defined as a binary measure, see **Supplementary Materials** for more details about phenotype definitions, antidepressant drugs included, and description of cohorts including genotype information.

Publicly available GWAS summary statistics were obtained from a GWAS on treatment response to antidepressants performed by the Psychiatric Genomics Consortium (PGC)^18^. We used summary statistics from the European sample of the genome-wide analysis of remission after antidepressant treatment (predominantly SSRIs) in individuals diagnosed with major depressive disorder (MDD). Summary statistics from two GWASs on antidepressant treatment response performed by the 23andMe Research Team from 23andMe, Inc.^20, 26^ were obtained upon request. In the GWASs from the 23andMe Research Team^20, 26^, treatment response and non-response to antidepressants was defined according to an antidepressant efficacy survey. We used separate summary statistics for treatment response to SSRIs and SNRIs. All GWAS samples and corresponding numbers of responders and non-responders are summarized in **Table 1**. All subjects provided written informed consent after receiving a complete description of the respective study.

**Table 1:** GWAS samples included in the GWAS meta-analysis of non-response to SSRIs, non-response to SNRIs, and non-response to SSRIs/SNRIs.

| GWAS sample | Antidepressant class | N total (Neff total) | N responders | N non-responders | Treatment response measure |
| --- | --- | --- | --- | --- | --- |
| Pain et al. 2022<sup>18</sup> | SSRIs | 5,151 (4,479) | 1,852 | 3,299 | Depression symptom scores |
| Li et al. 2016<sup>26</sup> +<br>Li et al. 2020<sup>20</sup> | SSRIs | 19,740 (17,586) | 13,130 | 6,610 | Antidepressant efficacy survey |
|  | SNRIs | 7,079 (6,943) | 4,030 | 3,049 |  |
| EstBB | SSRIs | 7,168 (4,272) | 5,862 | 1,306 | Antidepressant efficacy survey |
|  | SNRIs | 968 (584) | 789 | 179 |  |
| AGDS | SSRIs | 9,208 (6,902) | 6,908 | 2,300 | Antidepressant efficacy survey |
|  | SNRIs | 4,426 (4,304) | 2,580 | 1,846 |  |
| GLAD | SSRIs | 4,184 (2,416) | 3,452 | 732 | Antidepressant efficacy survey |
| UKB | SSRIs | 19,811 (10,632) | 16,648 | 3,163 | Antidepressant efficacy survey |
| deCODE | SSRIs | 49,062 (8,391) | 46,866 | 2,196 | Antidepressant switching |
|  | SNRIs | 8,674 (2,148) | 8,099 | 575 |  |
| Total | SSRIs | 114,324 (64,975) | 94,718 | 19,606 |  |
|  | SNRIs | 21,147 (16,560) | 15,498 | 5,649 |  |
|  | SSRIs/SNRIs | 135,471 (82,187) | 110,216 | 25,255 |  |

EstBB = Estonian Biobank, AGDS = Australian Genetics of Depression Study, GLAD = Genetic Links to Anxiety & Depression, UKB = UK Biobank

### Genome-wide meta-analyses

Meta-analyses of GWAS summary statistics were conducted using inverse-variance-weighted fixed effects models in METAL^27^. Separate meta-analyses were performed for non-response to SSRIs, non-response to SNRIs, and non-response to either SSRIs or SNRIs. To investigate if the effects of genome-wide significant SNPs were consistent across the datasets, we performed a heterogeneity test in METAL^27^. Due to differences in the definition of antidepressant treatment response across GWASs, we also performed sensitivity meta-analyses restricted to samples where treatment response was measured using only questionnaires (EstBB, AGDS, GLAD, UKB, and the GWASs from the 23andMe Research Team^20, 26^).

### Locus definition, variant annotation, and gene mapping

To define genetic loci based on the association summary statistics produced with METAL^27^, we used Functional Mapping and Annotation of GWAS (FUMA)^28^ with default settings. Genetic variants with a p-value <5e^-8^ and with a linkage disequilibrium (LD) r^2^ <0.6 with each other were defined as independent significant variants. Of these, variants with an LD r^2^ <0.1 were selected as lead variants. For a given lead variant, the borders of the genomic locus were defined as minimum/maximum positional coordinates over all corresponding candidate variants. Loci that were separated by less than 250 kb were then merged.

To investigate previous phenotype associations, we queried the identified loci in the GWAS catalogue^29^. SNPs were also queried for known expression quantitative trait loci (eQTLs) across multiple tissues using the GTEx portal (GTEx v8)^30^, as well as in different brain tissues using the BRAINEAC portal^31^. SNPs were annotated with Combined Annotation Dependent Depletion (CADD)^32^ scores, which predict how deleterious the SNP effect is on protein structure/function, and RegulomeDB^33^ scores, which predict the likelihood of regulatory functionality of SNPs.

The Open Targets Genetics platform (https://genetics.opentargets.org/)^34^ was used to map the identified loci to genes. This portal contains functional genomics data from different cell types and tissues retrieved from various repositories and datasets, including data on gene expression, chromatin conformation, and protein abundance that are aggregated to make robust connections between variants and likely causal genes. For each variant-gene prediction, a Variant to Gene (V2G) association score is provided to assign likely causal genes for a given variant. For each locus, we considered the top 3 genes with the highest V2G scores.

### Multi-trait conditional and joint analysis, SNP-based heritability, and genetic correlation

To account for the possible effect of major depression, we used multi-trait conditional and joint analysis (mtCOJO)^35^. We conditioned the effect of SNPs estimated for non-response to antidepressants on those of depression, using summary statistics of a GWAS on depression phenotypes^36^ including 246,363 cases and 561,190 controls, performed by the Psychiatric Genetics Consortium (PGC), excluding a 23andMe sample. This was done for non-response to SSRIs, non-response to SNRIs, and non-response to either SSRIs or SNRIs. We utilized linkage disequilibrium score regression (LDSC)^37^ to estimate the SNP-based heritability of our meta-analyzed GWAS as well as the GWAS summary statistics produced with mtCOJO. The SNP-based heritability was calculated on the observed scale. As non-response to antidepressants has been previously associated with genetics of other psychiatric traits as well as cognitive traits^38^, LDSC^37^ was used to estimate bivariate genetic correlations between antidepressant non-response and various psychiatric and cognitive traits, using summary statistics from the following GWASs: Alzheimer’s disease^39^, attention deficiency hyperactivity disorder (ADHD)^40^, autism spectrum disorder^41^, anxiety disorder^42^, bipolar disorder^43^, general cognitive performance^44^, educational attainment^45^, intelligence^46^, insomnia^47^, depression phenotypes^36^, mood instability^48^, neuroticism^49^, posttraumatic stress disorder (PTSD)^50^, schizophrenia^51^, subjective well-being^52^.

### Leave-One-Out Polygenic Scoring

Polygenic scores (PGSs) were constructed based on the association summary statistics produced in the GWAS meta-analysis of non-response to SSRIs, excluding each cohort in turn to create independent discovery and target datasets. The target samples were EstBB, UKB, AGDS, and deCODE. In EstBB, the PGS was calculated using the polygenic risk score continuous shrinkage (PRS-cs) approach^53^ with default options. In UKB and AGDS, PGSs were calculated using SBayesR^54^. In the deCODE sample, the PGS was calculated using LDPred^55^. In all four samples, the European sample of the 1000 Genomes Phase III^56^ was used to adjust for LD. To facilitate the interpretability of the results, PGSs were standardized within each sample (mean=0, SD=1) before statistical analysis. We performed logistic regression analyses to investigate if the PGS is associated with non-response to SSRIs in each of the four target samples. Age, sex, and the first ten principal components for genetic ancestry were included as covariates. Meta-analyses of results from the four cohorts were performed using the R-package metaplus^57^ with standard normal random effect. We also weighted the samples based on effective sample size, using the metafor^58^ R-package.

### Genetically informed drug prioritization

To estimate gene associations, we used GSA-MiXeR^59^, a novel gene-set analysis (GSA) tool that estimates fold enrichment and identifies gene-sets with greater biological specificity compared to standard GSA approaches^59^. We ran GSA-MiXeR for the summary statistics produced in the GWAS meta-analysis of non-response to SSRIs, SNRIs, and SSRIs/SNRIs. From the outputs, we chose genes with a positive MiXeR AIC value and an enrichment value of >10. All genes identified from GSA-MiXeR^59^ as well as Open Targets Genetics^34^ were then studied within networks of protein-protein interactions (PPIs) of gene products, using the latest version of the human protein interactome from the Barabási lab^60^, consisting of 18,217 unique proteins (nodes) interconnected by 329,506 PPIs after removing self-loops.

As most approved drugs do not target disease-associated proteins but bind to proteins in their network vicinity^61^, we defined a network not only including the genes identified from GSA-MiXeR^59^ and Open Targets Genetics^34^, but also genes in their immediate network proximity. To define antidepressant non-response networks (one for non-response to SSRIs, one for non-response to SNRIs, and one for non-response to SSRIs or SNRIs), we used the method network propagation^62-64^, implemented in the Cytoscape^65^ application Diffusion^64^. Genes identified from GSA-MiXeR^59^ and Open Targets Genetics^34^ were used as input query genes, and the top 1% of proteins from the diffusion output were included in the antidepressant non-response network. The Drug Gene Interaction Database (DGIdb, (https://www.dgidb.org/) v.5.0.6 (04/04/2024)^66^ was used to identify drug-gene interactions between approved drugs and genes in the three antidepressant non-response networks. Gene-set enrichment analysis (GSEA) was performed to test for enrichment of drug-gene interactions within our networks.

For the drugs interacting with genes in our networks, we retrieved drug-induced gene expression data (drug versus no drug) from the Connectivity Map (CMap) 2020^67, 68^, extracted from the Phase 2 data release of the Library of Integrated Cellular Signatures (LINCS) using the cmapR package^69^ in R version 4.3.1. As low drug concentrations in CMap have been shown to reduce the quality of the data^70^, we selected the highest concentration per drug.

We also performed transcriptome-wide association studies using S-PrediXcan^71^ to impute the genetically regulated gene expression using summary statistics produced in the GWAS meta-analysis of non-response to SSRIs, SNRIs, and SSRIs/SNRIs as input. Gene expression was imputed using high-performance gene expression prediction models trained using elastic net regression (downloaded from http://predictdb.org) trained on gene expression data from whole blood as well as 13 brain expression data sets from GTEx (version 8)^72, 73^ and covariance matrices calculated from 503 individuals with European ancestry from the 1000 Genomes project^56^. For gene expression in brain, S-MultiXcan^74^ was used to combine the S-PrediXcan results across the 13 brain tissues (more details in **Supplementary Methods**).

To evaluate if the drugs interacting with genes in our networks could change the predicted expression levels associated with antidepressant non-response (whether these drugs down-regulate genes up-regulated in antidepressant non-response or vice versa), the Spearman correlation ρ between the drug-induced gene expression perturbations and the predicted expression in drug target genes within the antidepressant non-response networks was calculated for each drug (separately for non-response to SSRIs, SNRIs, and SSRIs/SNRIs), where negative correlation coefficients indicate that the drug could reverse gene expression changes associated with antidepressant non-response.

## Results

### GWAS meta-analyses

From the meta-analysis of non-response to SSRIs, including a total of 114,324 individuals (19,606 non-responders and 94,718 responders), we identified one novel genome-wide significant locus (rs1106260 T/C; chr9: 138,111,032-138,136,174; OR = 1.0502; SE = 0.009; p-value = 3.55e-08). Another locus was found to be nominally significant (rs4884091 A/G; chr13: 78,971,895-79,003,053; OR = 1.0602, SE = 0.011, p-value = 6.38e-08). No genome-wide significant loci were identified from the meta-analysis of non-response to SNRIs, including 21,147 individuals (5,649 non-responders and 15,498 responders). However, one locus was nominally significant (rs10104815 T/C; chr8: 136,780,782-136,869,414; OR = 0.9213; p-value = 6.62e-08). From the meta-analysis of non-response to SSRIs/SNRIs, including a total of 135,471 individuals (25,255 non-responders and 110,216 responders), we identified one novel genome-wide significant locus (rs60847828 T/C; chr16: 8,460,781-8,490,789; OR = 1.0844; SE = 0.014; p-value = 1.18e-08), and one locus that was nominally significant (rs11677238 T/G; chr2: 114,336,733-114,512,514; OR = 1.0439; SE = 0.008; p-value = 5.424e-08). Manhattan plots from the three meta-analyses are shown in **Figure 1**.

**Figure 1:**
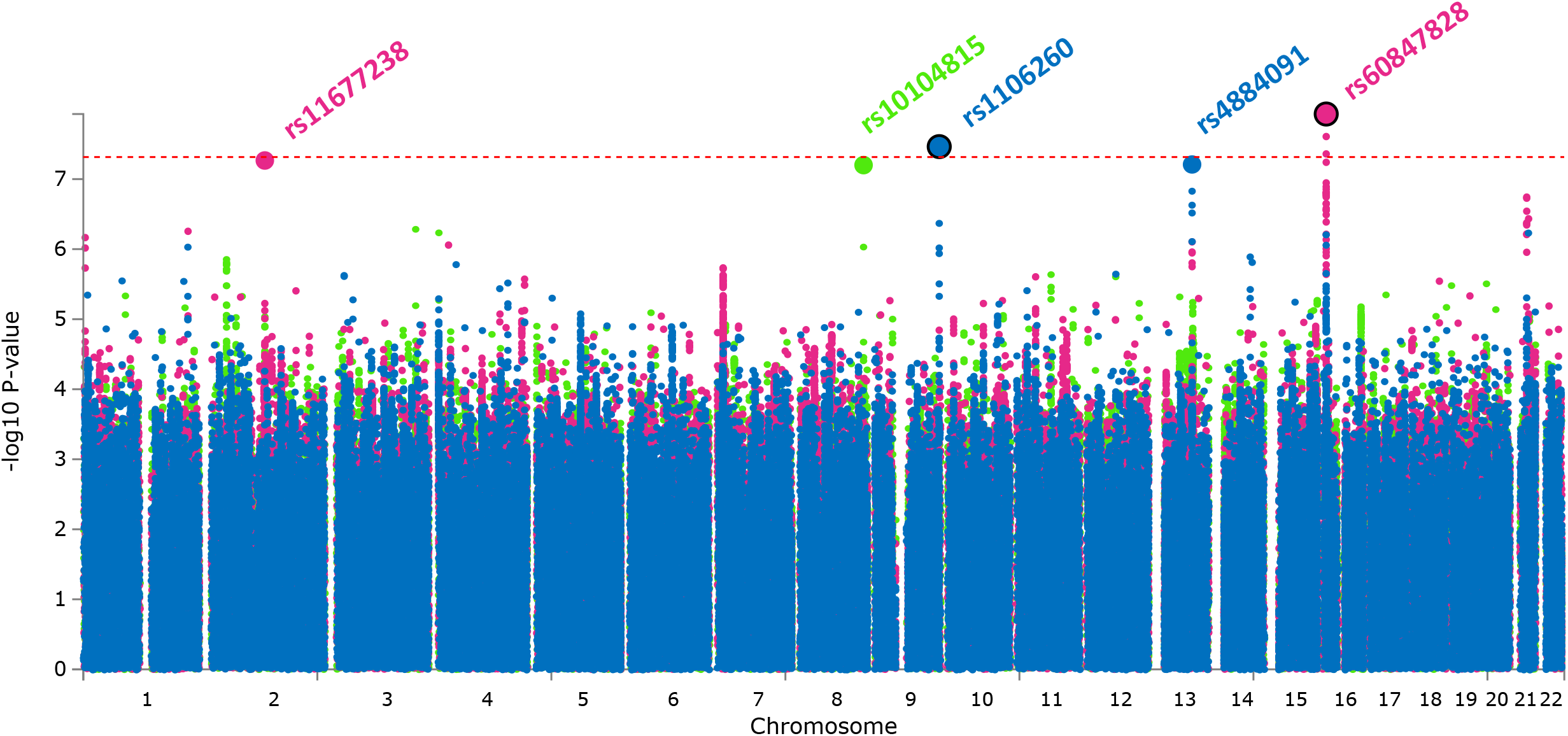
Manhattan plot showing genome-wide association results of the GWAS meta-analysis on non-response to SSRIs (blue), SNRIs (green), and SSRIs or SNRIs (pink). Genome-wide significant lead SNPs are encircled in black.

Quantile-quantile plots are shown in **Figure S1**. SNPs at the two genome-wide significant loci show no evidence of heterogeneity (p<5e-8), indicating that the effect is consistent across datasets (**Table S1, Figures S2-3**). When restricting the meta-analyses to samples where treatment response was measured using only questionnaires, similar results were obtained albeit the associations were no longer significant (p>5e-8) (**Figure S4-5**). All loci achieving genome-wide or nominally significance (p<1e-5) are reported in **Tables S2-7**.

Investigation of the genome-wide significant loci (rs1106260 and rs60847828) in the GWAS catalogue^29^ showed no previous associations. Functional annotation of rs1106260 and rs60847828 using FUMA^28^ does not suggest these SNPs to be deleterious (CADD scores <12.37) or likely to have regulatory functionality (RegulomeDB scores = 5-7). The top three genes with the highest V2G score for the identified locus for non-response to SSRIs (rs1106260) were *OLFM1, MRPS2*, and *PIERCE1*, of which the nearest gene is *OLFM1* (distance = 168,906 bp, downstream gene variant). No significant associations were found in the GTEx portal (GTEx v8)^30^ for the lead SNP (rs1106260). Additional assessment of the lead SNP (rs1106260) and gene expression of *OLFM1, MRPS2*, and *PIERCE1* in the BRAINEAC database^31^ showed significant associations between rs1106260 and gene expression of *OLFM1* in the medulla (p=0.015) and temporal cortex (p=0.005). The top three genes for the identified locus for non-response to SSRIs/SNRIs (rs60847828) were *TMEM114, METTL22*, and *ABAT*, of which the nearest gene is *TMEM114* (distance = 154,116 bp, intergenic variant). The lead SNP (rs60847828) was neither found in the GTEx portal (GTEx v8)^30^ nor in the BRAINEAC database^31^.

The locus that was nominally significant associated with SSRI non-response (rs4884091) has been previously associated with SSRI non-response in the GWAS from the 23andMe Team20. The top three genes with the highest V2G score for this locus were *OBI1, POU4F1*, and *EDNRB*, of which *POU4F1* is the nearest gene (260,067 bp, intron variant). One gene was mapped to the locus that was nominally significant associated with SNRI non-response (rs10104815), *KHDRBS3* (339,483 bp, intergenic variant), and this locus has been previously associated with non-response to SNRIs in the GWAS from the 23andMe Team20. The top three genes for the locus that was nominally significant associated with SSRI/SNRI non-response (rs11677238) were *SLC35F5, RABL2A*, and *PAX8*, of which the nearest gene is *RABL2A* (40,168 bp, upstream gene variant).

### Multi-trait conditional and joint analysis, SNP-based heritability, and genetic correlations

After conditioning on depression, the identified loci were still significantly associated with non-response to antidepressants (**Table S8-10**). SNP-based heritability estimates for all meta-analyses were in the range 0.019-0.028 and are reported in **Table S11**. Genetic correlation analyses show positive associations between non-response to antidepressants and most psychiatric traits, and negative associations with cognitive traits and subjective well-being (**Figure 2, Table S12-14**). Similar results were obtained using the meta-analyses restricted to questionnaire data (**Figure S6-8, Table S15-17**) and non-response to antidepressants conditioned on depression (**Figure S6-8, Table S18-20**).

**Figure 2:**
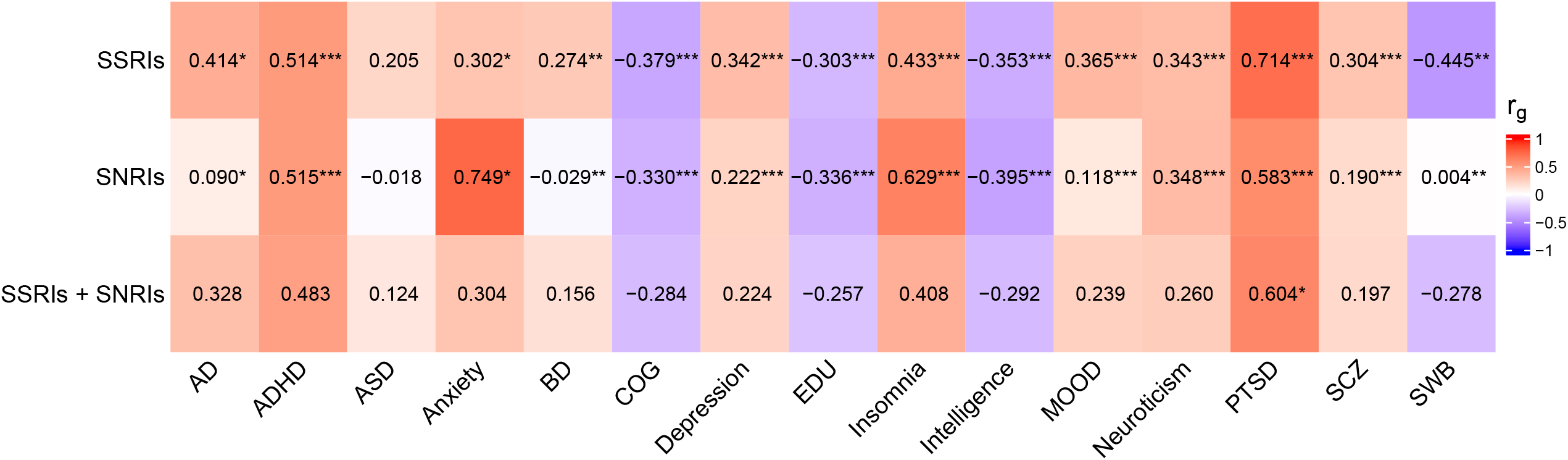
Genetic correlation between non-response to antidepressants and Alzheimer’s disease (AD), attention deficiency hyperactivity disorder (ADHD), autism spectrum disorder (ASD), anxiety disorder, bipolar disorder (BD), cognitive performance (COG), educational attainment (EDU), insomnia, intelligence, depression phenotypes, mood instability (MOOD), neuroticism, posttraumatic stress disorder (PTSD), schizophrenia (SCZ), subjective well-being (SWB). *p<0.05, **p<0.01, ***p<0.001

### Polygenic prediction of non-response to SSRIs

Meta-analysis of leave-one-out PGS analyses using SSRI non-response GWAS results in four samples showed a significant association with non-response to SSRIs (OR=1.016, CI=1.005-1.039, p-value=0.029), shown in **Figure 3**. However, in two out of the four samples, the PGS was not significantly associated with SSRI non-response (**Table S21**). Similar results were obtained when the samples were weighted based on effective sample size (OR=1.03, CI=1.00-1.05, p-value=0.025) (**Figure S9**).

**Figure 3:**
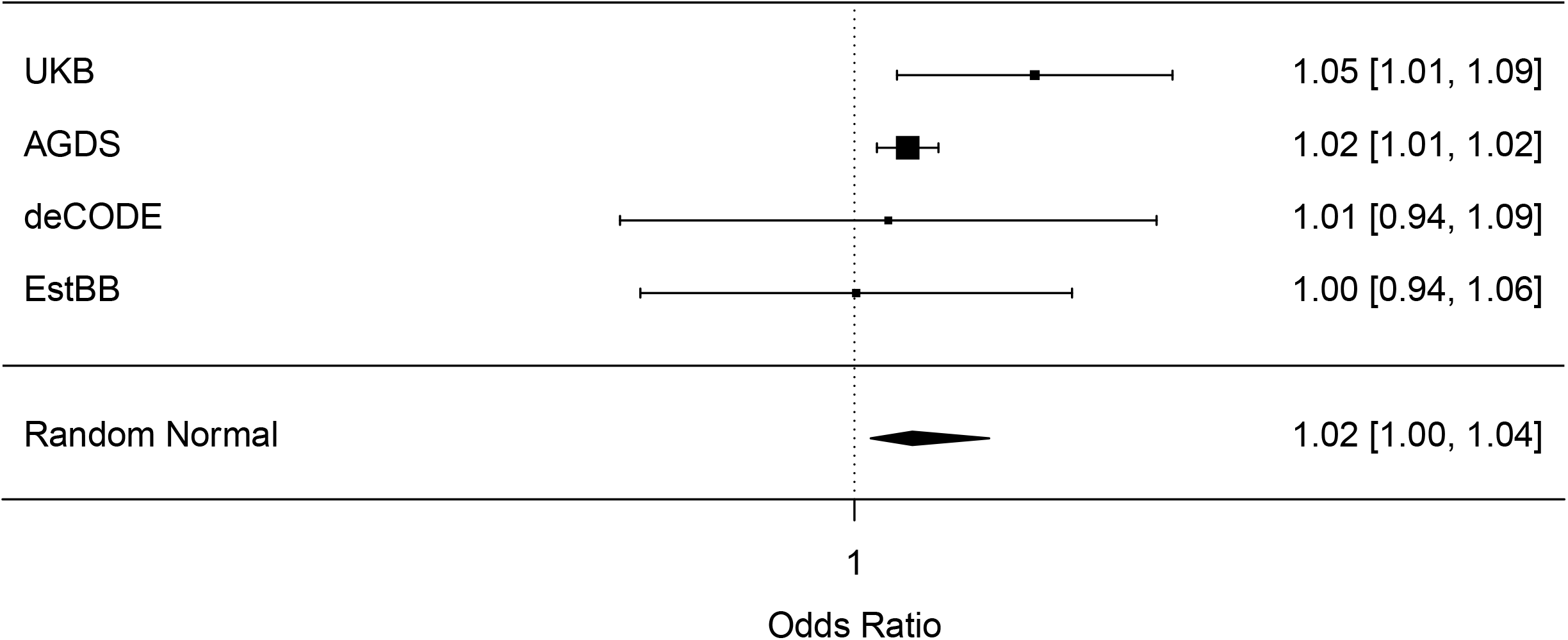
Forest plots showing the results from leave-one-out polygenic prediction of non-response to SSRIs in four independent cohorts, as well as meta-analyzed across these cohorts. Effects are reported as odds ratios (95% confidence interval).

### Genetically informed drug prioritization

From GSA-MiXeR, all genes with a positive AIC value and an enrichment score >10 can be found in **Table S22** (non-response to SSRIs, N=65), **Table S23** (non-response to SNRIs, N=108), and **Table S24** (non-response to SSRIs/SNRIs, N=76). The genes included in the three networks and the corresponding diffusion output values as well as their node degrees can be found in **Table S25** (SSRIs, N=252), **Table S26** (SNRIs, N=287), and **Table S27** (SSRIs/SNRIs, N=260).

Drug target genes in the SSRI non-response network were most significantly (p<5e-4) enriched for targets of the synthetic cannabinoid nabilone, and the target genes in the SSRI/SNRI non-response network were most significantly (p<5e-4) enriched for targets of bremelanotide, a drug developed to treat sexual dysfunction. However, after correction for the total number of drug-gene interactions (N=2,896), the enrichments remained non-significant (FDR>0.05). The drug target genes in the SNRI non-response network were significantly (FDR<0.05) enriched for several GABA receptor agonists (**Table S28-30**).

In the SSRI non-response network (**Figure S10**), six drugs (letrozole, clozapine, vandetanib, decamethonium, paclitaxel, budesonide) showed significant (p<0.05) opposite gene expression perturbations in drug (drug-induced expression) versus SSRI non-response-associated expression in drug target genes in brain tissue. For blood, eight drugs (temazepam, acetazolamide, chlordiazepoxide, ethionamide, amisulpride, rimonabant, clonazepam, fluorouracil) showed significant opposite gene expression (**Table S31-S34**). In the SNRI non-response network (**Figure S11**), two drugs (selegiline and norethindrone) showed significant (p<0.05) opposite gene expression perturbations in brain, and 2 drugs (dexamethasone and kinetin) in blood (**Table S35-38**). In the SSRI/SNRI non-response network (**Figure S12**), the drug simvastatin showed significant (p<0.05) opposite gene expression perturbations in brain, and the drug ascorbic acid showed significant opposite gene expression in blood (**Table S39-42**). However, after correction for multiple correlation analyses (number of drugs), all correlations remained non-significant (FDR>0.05). **Figure S13** summarizes the steps undertaken to identify drugs that could potentially address antidepressant non-response (more details in **Supplementary Results**), and the top drugs are shown in **Figure 4**.

**Figure 4:**
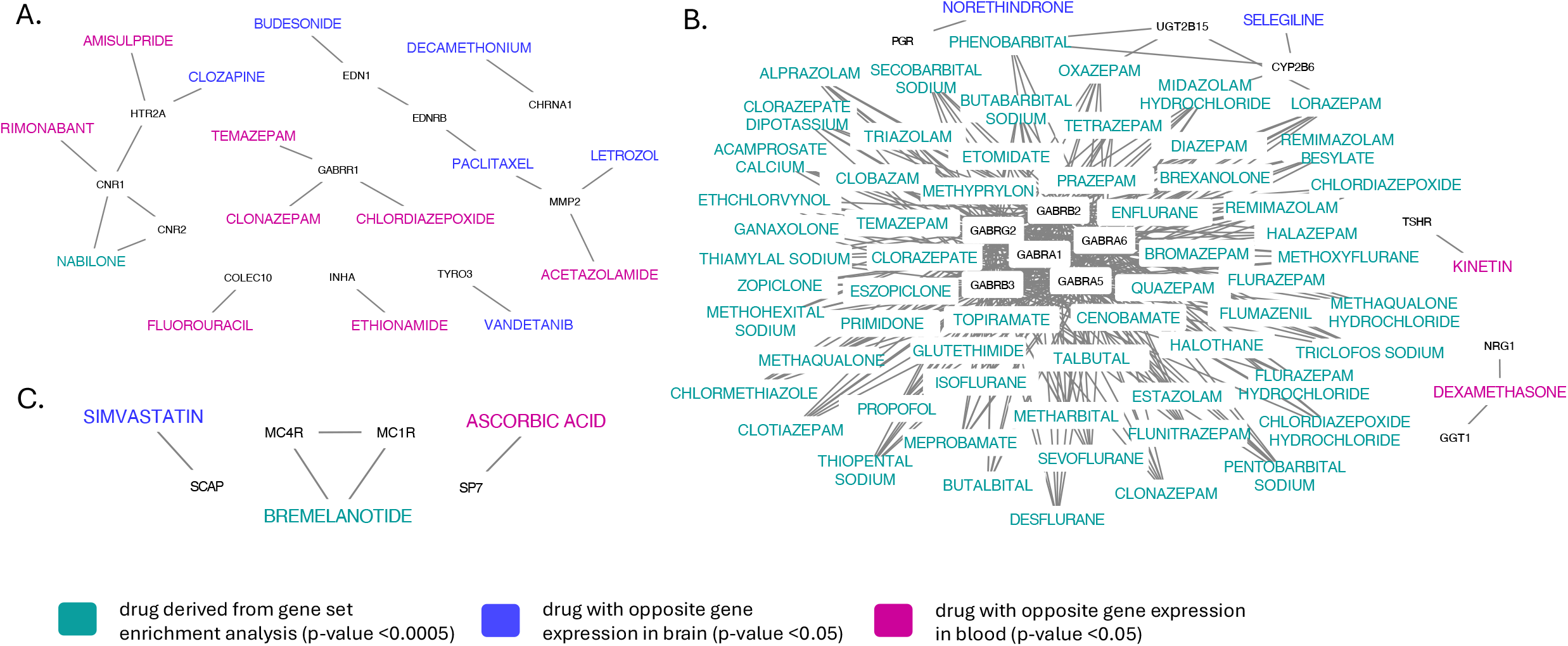
Top drugs identified based on gene-set enrichment analyses and drug-induced versus antidepressant non-response-associated gene expression, and their protein interaction partners in the SSRI non-response network (**A**), the SNRI non-response network (**B**), and SSRI/SNRI non-response network (**C**). Nodes refer to genes or drugs, and edges refer to gene-drug interactions or gene-gene interactions through identified protein-protein interactions between gene products (proteins).

## Discussion

In the present study, we identified two novel genome-wide significant loci associated with antidepressant non-response and showed that a polygenic score derived from our results predicted non-response to SSRIs in independent cohorts. By meta-analyzing real-world pharmacogenomic information on antidepressant non-response based on clinically assessed symptom scores, self-reported treatment outcomes, and data from eHR, this study represents the largest genetic investigation of non-response to antidepressants to date.

For the locus associated with non-response to SSRIs (rs1106260), the gene with the highest V2G score is *OLFM1*, which is also the nearest gene. The glycoprotein olfactomedin 1 (OLFM1) is highly expressed in the brain and participates in neural progenitor maintenance, cell death in brain, optic nerve arborization, and axonal growth^75, 76^. As OLFM1 plays a role in neuronal development, it has previously been suggested as a candidate gene for neuropsychiatric disorders^77^. In a study aiming to identify biomarkers for mood disorders, *OLFM1* showed strong evidence for predicting both depression and mania and was suggested as a target gene to treat depression^78^. One of the genes mapped to the identified locus for non-response to SSRIs/SNRIs (rs60847828) was *ABAT*. Variants within the GABA transaminase (*ABAT*) gene region have been associated with altered processing of somatosensory stimuli, indicating ABAT as a potential vulnerability marker for affective disorders^79^. Furthermore, it has been suggested that variants within *ABAT* affect valproic acid response^80^. Increasing evidence indicates that dysfunction of GABA, as well as glutamate systems contributes to depression-related behavior, and that ketamine’s antidepressant effects are related to its effect on glutamatergic and GABAergic neurons^81, 82^. Interestingly, our SNRI non-response network includes several GABA receptor genes, and we shortlist several drugs acting on the GABA system. These GABA receptor agonists may counteract the GABAergic deficits in depression^83^. We also shortlist several drugs with anti-inflammatory actions. A growing body of evidence supports an association between depression and inflammatory processes, and clinical trials have indicated antidepressant treatment effects for anti-inflammatory agents, both as add-on treatment and as monotherapy^84^. Our SSRI network includes *CNR1* and *CNR2*, the genes encoding the two main cannabinoid receptors, which are the primary targets for endogenous and exogenous cannabinoids. Studies suggest that the endocannabinoid system may be involved in the aetiology of depression and that targeting this system has the potential to relieve depressive symptoms^85^. However, the evidence that cannabinoids improve depressive disorders is weak and studies examining the effects of cannabinoids on mental disorders are needed^85, 86^.

Individual differences in pharmacological treatment response can often be attributed to genetic variability in cytochrome P450 genes (*CYP450*). In our antidepressant non-response GWASs as well as previous GWASs on antidepressant non-response, no association with CYP450 genes was detected. However, our SNRI network includes *CYP17A1* and *CYP2B6*, both identified from network propagation that prioritizes genes with biological and functional similarity to the input genes. Genetic variation in *CYP2B6* influences the metabolism of several SSRIs and SNRIs^87^. Guidelines from the Clinical Pharmacogenetics Implementation Consortium (CPIC) highlight the impact of *CYP2B6*, and *HTR2A* genotypes, among others, on antidepressant dosing, efficacy, and tolerability^87^. The pharmacodynamic gene *HTR2A* (serotonin-2A receptor) is included in our SSRI non-response network, also identified from network propagation.

We show an association between genetic liability of psychiatric disorders and non-response to antidepressants, which is in line with clinical studies^88^. We also identified a significant association between genetic propensity for cognitive phenotypes and improved antidepressant response. Similar genetic correlations have been shown in the previous GWAS on antidepressant response from the PGC^18^ as well as in a study investigating the genetic and clinical characteristics of treatment-resistant depression^38^. The strongest negative genetic correlations with non-response to antidepressants were observed for ADHD. This may indicate that phenotypic misspecification could underlie non-response to antidepressants. In fact, undetected ADHD has been associated with lack of response to SSRIs in MDD cases^89^. In adults, ADHD may be undiagnosed, and ADHD symptoms are often mistaken for those of their psychiatric comorbidities^90^.

Some limitations of the present study should be acknowledged. We combine various samples with differences in the assessment of treatment non-response, which introduces heterogeneity. Moreover, our study could potentially include individuals who were treated with antidepressants for conditions other than depression, especially in the sample where eHR were used to define non-response based on switching as proxy phenotype. Heterogeneity could also be introduced by differences in dosing, treatment duration, and co-treatment with other drugs. However, we performed heterogeneity tests and restricted our meta-analyses to samples where non-response was measured using only similar questionnaires, and these sensitivity analyses showed that the results were consistent across samples. It should also be noted that the individuals in our samples are of European ancestry, and our results may therefore not be directly translatable to other ethnicities.

In conclusion, these results suggest that meta-analyses of GWAS utilizing real-world measures of treatment outcomes can increase sample sizes to improve the discovery of variants associated with non-response to antidepressants.

## Data Availability

Data produced in the present study are available upon reasonable request to the authors.

## Author contributions

EK, OAA, and KSO conceived the study and were involved in study design. EK, TJ, GE, BM, AH, LMGM, KK, YL, YX, and KSO conducted analyses. EK drafted the initial manuscript. All authors contributed to data interpretation and editing of the manuscript.

## Acknowledgements

We would like to thank the research participants and employees of 23andMe, Inc. for making this work possible. This work was partly performed on the TSD (Tjeneste for Sensitive Data) facilities, owned by the University of Oslo, operated and developed by the TSD service group at the University of Oslo, IT-Department (USIT). Computations were also performed on resources provided by UNINETT Sigma2—the National Infrastructure for High Performance Computing and Data Storage in Norway (NS9666S). Computations of the UKB data were enabled by resources in project sens2017519 provided by the National Academic Infrastructure for Supercomputing in Sweden (NAISS) at UPPMAX, funded by the Swedish Research Council through grant agreement no. 2022-06725. We gratefully acknowledge support from the Research Council of Norway (RCN) (296030, 223273, and 334920) and from the National Institutes of Health (NIH 5R01MH124839-02). This project has received funding from the European Union’s Horizon 2020 research and innovation programme under grant agreement No 964874 (REALMENT), as well as from the US NIMH (grant R01 MH123724) and from the European Union’s Horizon 2021 research project Psych-STRATA (EU HORIZON-HLTH-2021, grant 101057454). The AGDS was primarily funded by National Health and Medical Research Council (NHMRC) of Australia grant 1086683, with additional funding from MRFF1200644 and MRFF2024891. BLM and SEM were supported by Australian NHMRC Investigator grants (APP2017176 and APP1172917 respectively). YL was supported by the European Research Council grant (grant agreement No: 101042183) and the Swedish Research Council grant (2021-02615_VR). MER thanks the support of the Rebecca L Cooper Medical Research Foundation through an Al & Val Rosenstrauss Fellowship (F20231230). GB, TH, and YL declare that this study represents independent research part-funded by the National Institute for Health and Social Care Research (NIHR) Biomedical Research Centre at South London and Maudsley NHS Foundation Trust and King’s College London. The views expressed are those of the author(s) and not necessarily those of the UK National Health Service, the NIHR or the Department of Health. We gratefully acknowledge the participation of all South London and Maudsley NIHR BioResource volunteers within the National NIHR BioResource and thank the South London and Maudsley BioResource staff for their help with volunteer recruitment.

