## Supplementary_material for "Genome-wide meta-analyses of non-response to antidepressants identify novel loci and potential drugs"

### Supplementary Methods

#### GWAS sample description

##### *Estonian Biobank*

Estonian Biobank samples were genotyped in Core Genotyping Lab of Institute of Genomics, University of Tartu using Illumina GSAv1.0, GSAv2.0, GSAv2.0_EST, and GSAv3.0_EST arrays and Illumina GenomeStudio v2.0.4. During the quality control all individuals with call-rate < 95% or mismatch between genetic sex and sex in the phenotype data, were excluded from the analysis. Variants were filtered by call rate < 95%, MAF < 1% and HWE p-value < 1e-4. The total number of SNPs that passed all filters and were used for phasing is 308,302. Reference-free phasing was performed using Eagle v2.4.1[1] with original Eagle hg19 recombination map and default settings. The total number of SNPs used for the imputation was 296,903. Imputation was performed with Beagle 5.4[2] and default settings, using the Estonian population specific imputation reference of 2056 WGS samples. Principal component analysis (PCA) was performed in PLINK 2[3] using quality controlled and pruned genotyped SNPs.

Treatment response and non-response was defined according to data from the Estonian Biobank mental health online survey. Individuals were asked “Was the prescribed medication effective?”. Individuals who said they have been prescribed medications for more than 2 weeks and answered “No” were coded as non-responders, and those who have been prescribed medications for more than 2 weeks and answered “Yes” were coded as responders. SSRIs included escitalopram, sertraline, fluoxetine, citalopram, and paroxetine. SNRIs included duloxetine and venlafaxine. GWAS on antidepressant non-response was performed with REGENIE 3.2[4], separately for SSRIs (1,306 non-responders and 5,862 responders) and SNRIs (179 non-responders and 789 responders).

##### *Australian Genetics of Depression Study*

In the Australian Genetics of Depression Study (AGDS)[5], genotyping was conducted using the Illumina Global Screening Array V.2.0 (GSA). GSA was developed by human genetic disease researchers to maximize utility for gene mapping. It includes a common variant backbone component that maximizes information for imputation of common variants in multiple ethnic populations as well as a suite of common and rare variants selected for known or likely association with a range of genetic disorders. SNPs with MAF >0.01 and INFO >0.3 were included.

Treatment response and non-response was defined according to questionnaire data. Individuals were asked “How well does/did antidepressant [X] work for you?” and got the following four possible answers to choose from: “Not at all well”, “Moderately well”, “Very well”, “Don’t know”. Individuals were also asked “Why were you prescribed [X]?”. Those who answered that they were prescribed antidepressants for depression were included. Individuals who answered “Not at all well” to at least one antidepressant were coded as non-responders, and individuals who answered “Very well” or “Moderately well” to all antidepressants were coded as responders. SSRIs included sertraline, escitalopram, citalopram, fluoxetine, and paroxetine. SNRIs included venlafaxine, desvenlafaxine, and duloxetine. GWAS on antidepressant non-response was performed with SAIGE v0.44 including age, sex, and ten principal components as covariates, which was done separately for SSRIs (2,300 non-responders and 6,908 responders) and SNRIs (1,846 non-responders and 2,580 responders).

***Genetic Links to Anxiety & Depression Study***

The samples of the Genetic Links to Anxiety & Depression (GLAD) Study[6] were genotyped as part of core NIHR BioResource funding. For genotyping, the UK Biobank v2 Axiom array was used, consisting of >850,000 genetic variants[7], designed to give optimal information about other correlated genetic variants. Imputation of the data with large whole genome sequencing reference datasets yields >10 million common genetic variants per individual. We use Affymetrix software and the UK Biobank pipeline software to assign genotypes and perform standard quality control measures in PLINK[3].

Treatment response and non-response was defined according to questionnaire data. Individuals were asked "How well does/did antidepressant [X] work for you?" and got the following four possible answers to choose from: "Not at all well", "Moderately well", "Very well", "Don't know". Individuals were also asked "Why were you prescribed [X]?". Those who answered that they were prescribed antidepressants for depression were included. Individuals who answered "Not at all well" to at least one antidepressant were coded as non-responders, and individuals who answered "Very well" or "Moderately well" to all antidepressants were coded as responders. We performed a GWAS on non-response to SSRIs (3,452 responders and 732 non-responders).

***UK Biobank***

The UK Biobank has ethical approval from the NHS National Research Ethics Service as a research tissue bank (References 16/NW/0274 and 11/NW/0382) and all necessary patient/participant consent was obtained. DNA was extracted from stored blood samples, and genotyp- ing was performed using 2 closely related arrays with over 95% common content, the UK BiLEVE array and the UK Biobank Axiom array. Samples were analyzed in batches of approximately 4700 items, initial quality control of the genotyping data was done by Affymetrix. More detailed information on the array and sample processing is available on the UK Biobank website (https://www.ukbiobank.ac.uk). Quality control and imputation (to over 90 million SNPs, indels and large structural variants) has been performed by a collaborative group headed by the Wellcome Trust Centre for Human Genetics. Post-imputation quality control (QC) was performed based on genotype call rate <10%, minor allele frequency (MAF) <1%, and SNP missingness <5%. For details on genotyping see Bycroft et al. 2018[7]. Individuals with non-European ancestry were excluded for the present analysis.

From the UK Biobank (UKB)[8], we used the mental well-being online question "Have you ever tried any of the following medications for at least two weeks?" (Field ID 29039 from category 1502, depression), from which we included individuals who reported having tried SSRIs. Individuals were also asked "Has the prescribed medication helped you feel better?" with the following four options: "Yes, at least a little", "No", "Do not know", "Prefer not to answer". Individuals who answered "Yes, at least a little" were classified as responders, and those who answered "No" were classified as non-responders, and excluded those who did not know or did not want to answer. SSRIs included citalopram, fluoxetine, sertraline, and paroxetine. We performed a GWAS on non-response to SSRIs (16,648 responders and 3,163 non-responders) using Genome-wide Complex Trait Analysis (GCTA)[9] version v1.94.1, adding age, sex, and the first 10 principal components as covariates. UKB data use was approved via application 22224.

***deCODE Genetics***

At deCODE Genetics, genetic variants identified in the whole-genome sequences of 64,460 individuals were imputed into the datasets of 173,025 chip-genotyped participants using long-range phasing[10]. SNP calling was performed with Graphtyper[11-13]. Sequencing was conducted with Illumina's GAIIx, HiSeq, HiSeqX, and NovaSeq technologies.

We utilized prescription registry data from the deCODE genetics Icelandic cohort (prescribed medications for Icelanders from 2003-2019). These records were sourced from Landlaeknir, the national directorate of health. Approval for the study of depression was granted by the Icelandic Data Protection Authority and the National Bioethics Committee of Iceland (Approval no: VSNb2016040011/03.01). We defined switching phenotypes, i.e., individuals that switch from one antidepressant drug to another antidepressant drug, and individuals who continued being prescribed the same antidepressant. We excluded all prescriptions where the individual was older than 60 years, based on year of birth rounded to the nearest year divisible by 5. Individuals who switched from the first prescription of a specific antidepressant (SSRI or SNRI) to another antidepressant (any) between 5-35 days and who had max 1 additional prescription of the specific antidepressant after the switch were classified as non-responders. Individuals who had at least three prescriptions of the same antidepressant in a 90-day window were classified as responders. SSRIs included escitalopram, sertraline, fluoxetine, citalopram, and paroxetine. SNRIs included duloxetine and venlafaxine. GWAS on antidepressant non-response was performed separately for SSRIs (2,196 non-responders and 46,866 responders) and SNRIs (575 non-responders and 2,196 responders). We tested the association between sequence variants and drug switching phenotypes using logistic regression with an additive model and software from deCODE genetics[11]. To control for inflation from cryptic relatedness and population stratification, we applied LD-score regression[14]. Adjustments were made for sex, county of origin, current age or age at death (including first and second order terms), genotype availability, and an indicator for the individual's lifetime overlapping with the phenotype collection period.

***Publicly available GWAS summary statistics***

Publicly available GWAS summary statistics were obtained from the following GWASs: A GWAS on treatment response to antidepressants performed by the Psychiatric Genomics Consortium (PGC)[15], and two GWASs on antidepressant treatment response performed by the 23andMe Research Team[16, 17]. From the GWAS from the PGC[15], we used summary statistics from the European sample of the genome-wide analysis of remission after antidepressant treatment (predominantly SSRIs) in MDD patients ($N_{remit}$ = 1,852, $N_{nonremit}$ = 3,299). Remission was defined as a binary measure attained when a patient's depression symptom score decreased to pre-specified thresholds for symptom rating scales. Remission thresholds for scales were: Montgomery And Åsberg Depression Rating Scale (MADRS)[18] ≤ 10, Quick Inventory of Depressive Symptomatology (QIDSC)[19] ≤ 5, Hamilton Depression Rating Scale[20] (HAMD-17) ≤ 7, HAMD-21 ≤ 7, and Beck Depression Inventory (BDI)[21] ≤ 9. Patients who did not reach these thresholds were classified as non-remitting. In the GWASs from the 23andMe Research Team[16, 17], treatment response to antidepressants was defined according to an antidepressant efficacy survey. Responders included individuals treated with only 1 antidepressant for ≥ 3 weeks and answered ‘Yes’ to; overall treatment effect = ´helpful´ or ´very helpful´. Non-responders included individuals treated with ≥ 2 antidepressant for ≥ 5 weeks and answered ‘No’ to; overall treatment effect = ´helpful´ or ´very helpful´. We used the separate summary statistics for treatment response on SSRIs and serotonin-norepinephrine reuptake inhibitors (SNRIs). In the GWAS published in 2016[17], these included 6,348 SSRI responders and 3,340 SSRI non-responders, and 2,547 SNRI responders and 1,567 SNRI non-responders. In the GWAS published in 2020[16], these included 8,491 SSRI responders and 4,046 SSRI non-responders, and 2,055 SNRI responders and 1,950 SNRI non-responders.

**Genetically informed drug prioritization**

To estimate gene associations, we used GSA-MiXeR[22], a novel gene-set analysis (GSA) tool that estimates fold enrichment and identifies gene-sets with greater biological specificity compared to standard GSA approaches[22]. We ran GSA-MiXeR for the summary statistics produced in the GWAS meta-analysis of non-response to SSRIs, non-response to SNRIs, and non-response to SSRIs/SNRIs. From the outputs, we chose genes that had a positive MiXeR AIC value and an enrichment value of >10. All genes identified from GSA-MiXeR[22] as well as Open Targets Genetics[23] were then studied within networks of protein-protein interactions (PPIs) of gene products, using the latest version of the human protein interactome from the Barabási lab[24], consisting of 18,217 unique proteins (nodes) interconnected by 329,506 PPIs (edges or links) after removing self-loops.

As most approved drugs do not target disease-associated proteins but bind to proteins in their network vicinity[25], we defined a network not only including the genes identified from GSA-MiXeR[22] and Open Targets Genetics[23], but also genes in their immediate network proximity. To define antidepressant non-response networks (one for non-response to SSRIs, one for non-response to SNRIs, and one for non-response to SSRIs or SNRIs), we used the method network propagation[26-28], implemented in the Cytoscape[29] application Diffusion[28]. Starting with a chosen set of input proteins, information from their PPIs is transferred to all other proteins in the interactome and received from them through an iterative process. Network proximity between proteins is scored depending on their PPIs, where higher diffusion output values relate to higher relatedness to the input proteins[26-28]. Genes identified from GSA-MiXeR[22] and Open Targets Genetics[23] were used as input query genes, and the top 1% of proteins from the diffusion output were included in the antidepressant non-response network. This was done separately for non-response to SSRIs, non-response to SNRIs, and non-response to SSRIs/SNRIs.

The Drug Gene Interaction Database (DGIdb, (https://www.dgidb.org/) v.5.0.6 (04/04/2024)[30] was used to identify drug-gene interactions between approved drugs and genes in the three antidepressant non-response networks. The DGIdb provides information on drug-gene interactions from 28 diverse sources that are aggregated and normalized. The database collects drug-gene interactions based on information about therapeutic targets and their corresponding drugs, knowledge from clinical trials, as well as potentially clinically actionable drug-gene associations based on metadata such as molecule structure and molecular weight[30]. Gene-set enrichment analysis (GSEA) was performed to test for enrichment of drug-gene interactions within our networks.

For the drugs interacting with genes in our networks, we retrieved drug-induced gene expression data (drug versus no drug) from the Connectivity Map (CMap) 2020[31, 32], extracted from the Phase 2 data release of the Library of Integrated Cellular Signatures (LINCS) (level5_beta_trt_cp_n720216x12328.gctx.gz available at https://clue.io/data/CMap2020#LINCS2020) using the cmapR package[33] in R version 4.3.1. As low drug concentrations in CMap have been shown to reduce the quality of the data[34], we selected the highest concentration per drug.

We also performed transcriptome-wide association studies using S-PrediXcan[35] to impute the genetically regulated gene expression using summary statistics from GWAS. S-PrediXcan first predicts gene expression levels based on reference transcriptome data and then estimates the correlation between the gene expression levels and a phenotype using GWAS summary statistics[35]. For S-PrediXcan, we used the summary statistics produced in the GWAS meta-analysis of non-response to SSRIs, non-response to SNRIs, and non-response to SSRIs/SNRIs as input. Gene expression was imputed using high-performance gene expression prediction models trained using elastic net regression (downloaded from http://predictdb.org) trained on gene expression data from whole blood as well as 13 brain expression data sets from GTEx (version 8)[36, 37] and covariance matrices calculated from 503 individuals with European ancestry from the 1000 Genomes project[38]. For gene expression in brain, S-MultiXcan[39] was used to combine the S-PrediXcan results across the 13 brain tissues.

To evaluate if the drugs interacting with genes in our networks could change the predicted expression levels associated with antidepressant non-response (whether these drugs down-regulate genes up-regulated in antidepressant non-response or vice versa), the Spearman correlation ρ between the drug-induced gene expression perturbations and the predicted expression in drug target genes within the antidepressant non-response networks was calculated for each drug (separately for non-response to SSRIs, non-response to SNRIs, and non-response to SSRIs/SNRIs), where negative correlation coefficients indicate that the drug could reverse gene expression changes associated with antidepressant non-response.

### Supplementary Results

##### Genetically informed drug prioritization

From GSA-MiXeR, all genes with a positive AIC value and an enrichment score >10 can be found in **Table S22** (non-response to SSRIs, N=65), **Table S23** (non-response to SNRIs, N=108), and **Table S24** (non-response to SSRIs/SNRIs, N=76). All SSRI non-response genes (N=65) were present in the human protein interactome. Of the 108 SNRIs non-response genes, 105 were present in the human protein interactome. Of the 76 SSRIs/SNRIs non-response genes, 74 were present in the human protein interactome. To define antidepressant non-response networks (one for non-response to SSRIs, one for non-response to SNRIs, and one for non-response to SSRIs/SNRIs), we performed three network propagation[26-28] analyses using the GSA-MiXeR genes and the genes from Open Target Genetics (six genes for non-response to SSRIs, one gene for non-response to SNRIs, and six genes for non-response to SSRIs/SNRIs) as input genes, and chose the top 1% of genes from the diffusion output. In total, 252 genes were included in the SSRI non-response network, 287 genes in the SNRI non-response network, and 260 genes were included in the SSRI/SNRI non-response network. The genes included in the three networks and the corresponding diffusion output values as well as their node degrees can be found in **Table S25** (SSRIs), **Table S26** (SNRIs), and **Table S27** (SSRIs/SNRIs).

Of the 252 genes in the SSRI non-response network, 38 interact with 230 drugs with regulatory approval. Of the 287 genes in SNRI non-response network, 50 interact with 414 drugs. Of the 260 genes in SSRI/SNRI non-response network, 32 interact with 158 drugs. We performed GSEA to test for the enrichment of these drug-gene interactions. The 38 drug target genes in the SSRI non-response network were most significantly ($p<0.0005$) enriched for targets of the synthetic cannabinoid nabilone, and the 32 target genes in the SSRI/SNRI non-response network were most significantly ($p<0.0005$) enriched for targets of bremelanotide, a drug developed to treat sexual dysfunction. However, after correction for the total number of drug-gene interactions (N=2,896), the enrichments remained non-significant (FDR>0.05). The 50 drug target genes in the SNRI non-response network were significantly (FDR<0.05) enriched for several GABA receptor agonists (**Table S28-30**).

Of the 252 genes in the SSRI non-response network (**Figure S10**), SSRI non-response-associated gene expression could be imputed for 156 and 83 genes in brain and blood, respectively (**Table S31-32**). Of the 38 drug target genes in the SSRI-response network (**Figure S10**), SSRI non-response-associated gene expression could be imputed for 21 and 10 genes in brain and blood, respectively. Of these 21 and 10 genes, 19 and 9 were present in CMap, respectively. Of the 230 drugs in the SSRI-response network, 94 are present in CMap. Of the 94 drugs, 6 drugs (letrozole, clozapine, vandetanib, decamethonium, paclitaxel, budesonide) showed significant ($p<0.05$) opposite gene expression perturbations in drug (drug-induced expression) versus SSRI non-response-associated expression in brain in the 19 drug target genes in the SSRI non-response network (**Table S33**). For blood, 8 drugs (temazepam, acetazolamide, chlordiazepoxide, ethionamide, amisulpride, rimonabant, clonazepam, fluorouracil) showed significant opposite gene expression in the 9 drug target genes (**Table S34**).

Of the 287 genes in SNRI non-response network (**Figure S11**), SNRI non-response-associated gene expression could be imputed for 186 and 19 genes in brain and blood, respectively (**Table S35-36**). Of the 50 drug target genes in the SNRI-response network (**Figure S11**), SNRI non-response-associated gene expression could be imputed for 36 and 14 genes in brain and blood, respectively. Of these 36 and 14 genes, 35 and 14 were present in CMap, respectively. Of the 414 drugs in the SNRI-response network, 200 are present in CMap. Of these 200 drugs, 2 drugs (selegiline and norethindrone) showed significant ($p<0.05$) opposite gene expression perturbations in drug (drug-induced expression) versus SNRI non-response-associated expression in brain in the 35 drug target genes in the SNRI non-response network (**Table S37**). For blood, 2 drugs (dexamethasone and kinetin) showed significant opposite gene expression in the 14 drug target genes (**Table S38**).

Of the 260 genes in SSRI/SNRI non-response network (**Figure S12**), SSRI/SNRI non-response-associated gene expression could be imputed for 153 and 88 genes in brain and blood, respectively (**Table S39-40**). Of the 32 drug target genes in the SSRI/SNRI-response network (**Figure S12**), SSRI/SNRI non-response-associated gene expression could be imputed for 21 and

11 genes in brain and blood, respectively. Of these 21 and 11 genes, 19 and 11 were present in CMap, respectively. Out of the 158 drugs in the SSRI/SNRI network, 70 are present in CMap. Of these 70 drugs, 1 drug (simvastatin) showed significant ($p<0.05$) opposite gene expression perturbations in drug (drug-induced expression) versus SSRI/SNRI non-response-associated expression in brain in the 19 drug target genes in the SSRI/SNRI non-response network (**Table S41**). For blood, 1 drug (ascorbic acid) showed significant opposite gene expression in the 11 drug target genes (**Table S42**). However, after correction for multiple correlation analyses (number of drugs), all correlations remained non-significant (FDR>0.05).

#### Supplementary Figures

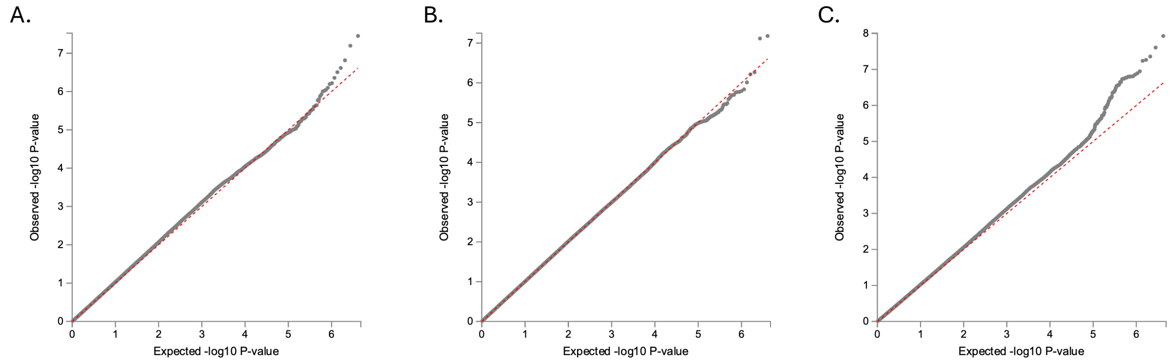


**Figure S1**: Quantile-quantile plots of the GWAS meta-analysis on non-response to SSRIs (**A**), non-response to SNRIs (**B**), and non-response to SSRIs or SNRIs (**C**).

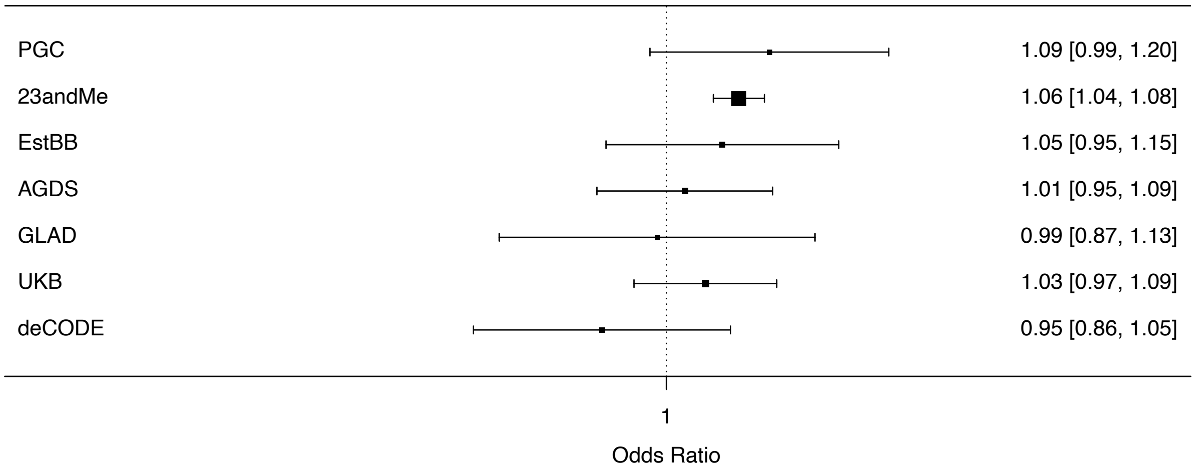


**Figure S2**: Forest plot showing the association with non-response to SSRIs for rs1106260 (T>C) in the different samples.

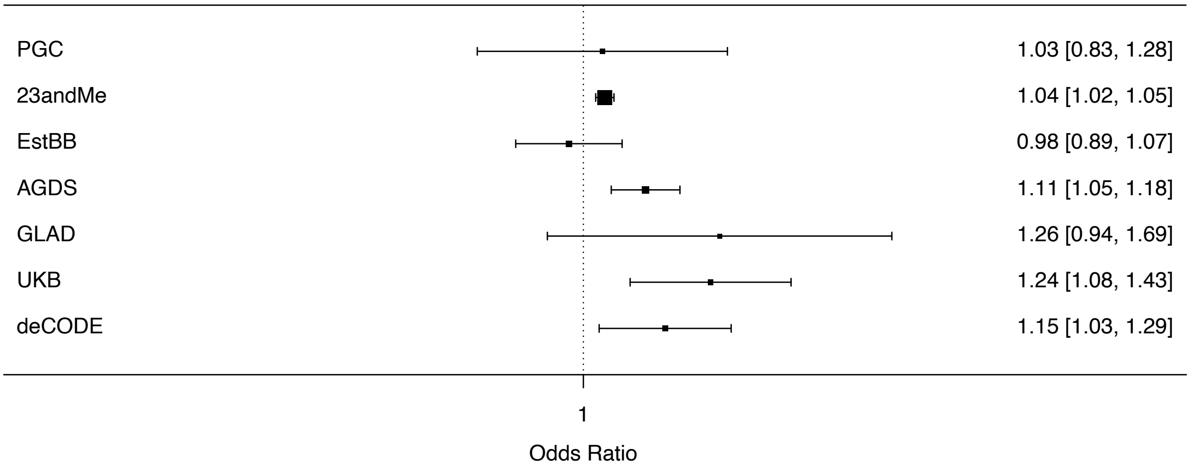


**Figure S3**: Forest plot showing the association with non-response to SSRIs/SNRIs for rs60847828 (T>C) in the different samples.

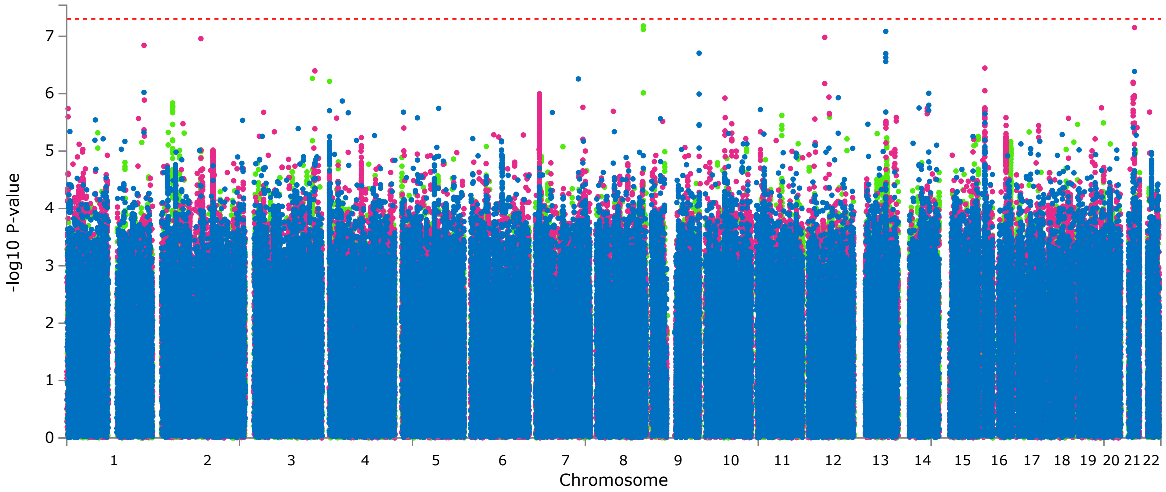


**Figure S4**: Manhattan plot showing genome-wide association results of the GWAS meta-analysis on non-response to SSRIs (blue), non-response to SNRIs (green), and non-response to SSRIs or SNRIs (pink), restricted to samples where treatment response was measured using only questionnaires.

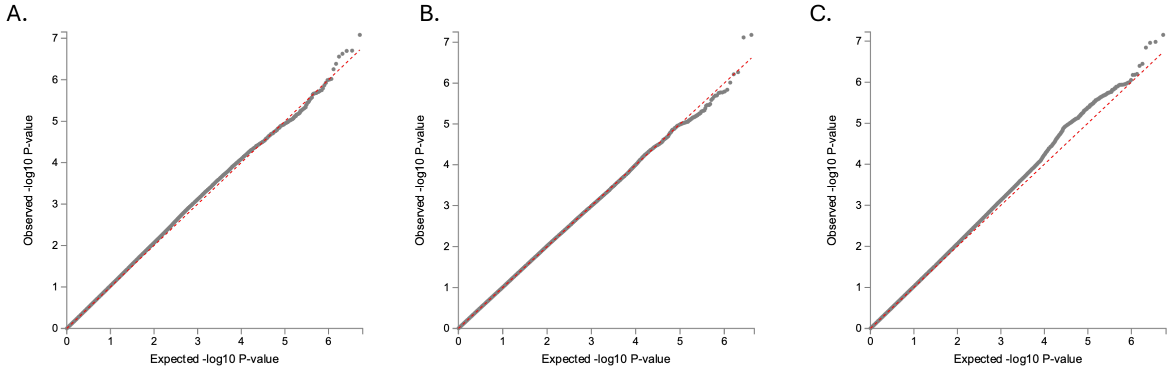


**Figure S5**: Quantile-quantile plots of the GWAS meta-analysis on non-response to SSRIs (**A**), non-response to SNRIs (**B**), and non-response to SSRIs or SNRIs (**C**), restricted to samples where treatment response was measured using only questionnaires.

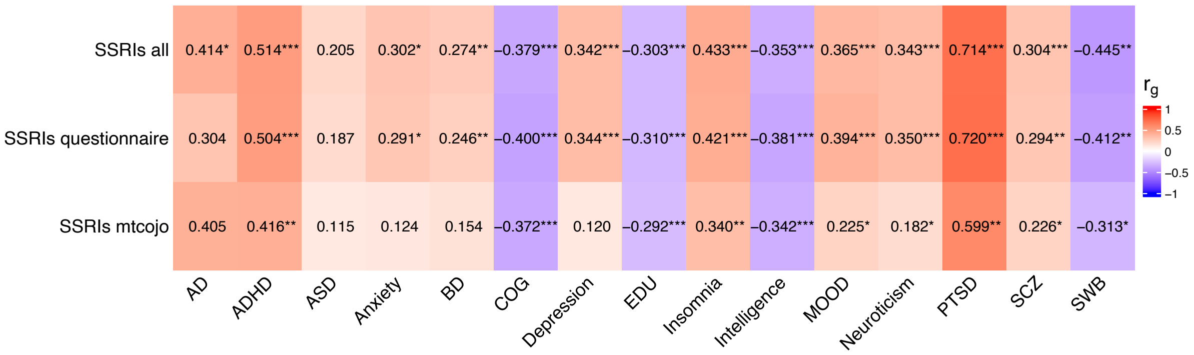


**Figure S6**: Genetic correlation between non-response SSRIs (from various meta-analyses) and Alzheimer´s disease (AD), attention deficiency hyperactivity disorder (ADHD), autism spectrum disorder (ASD), anxiety disorder, bipolar disorder (BD), cognitive performance (COG), educational attainment (EDU), insomnia, intelligence, depression phenotypes, mood instability (MOOD), neuroticism, posttraumatic stress disorder (PTSD), schizophrenia (SCZ), subjective well-being (SWB). *p<0.05, **p<0.01, ***p<0.001

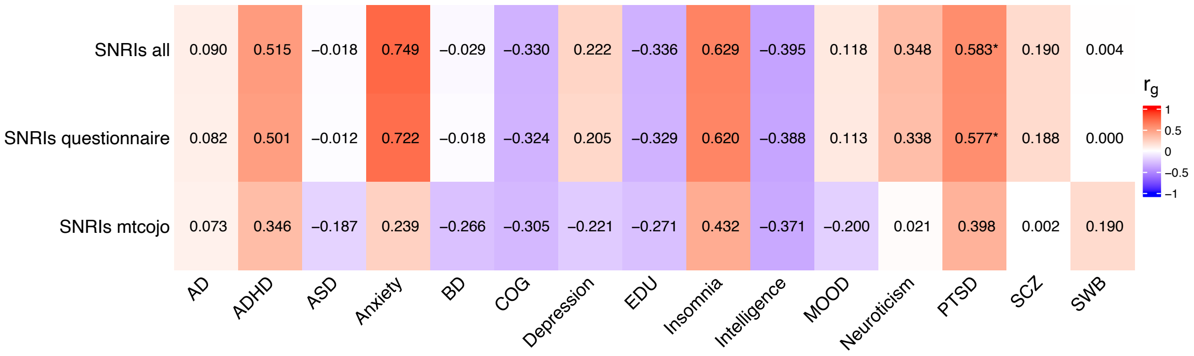


**Figure S7**: Genetic correlation between non-response SNRIs (from various meta-analyses) and Alzheimer´s disease (AD), attention deficiency hyperactivity disorder (ADHD), autism spectrum disorder (ASD), anxiety disorder, bipolar disorder (BD), cognitive performance (COG), educational attainment (EDU), insomnia, intelligence, depression phenotypes, mood instability (MOOD), neuroticism, posttraumatic stress disorder (PTSD), schizophrenia (SCZ), subjective well-being (SWB). *p<0.05, **p<0.01, ***p<0.001

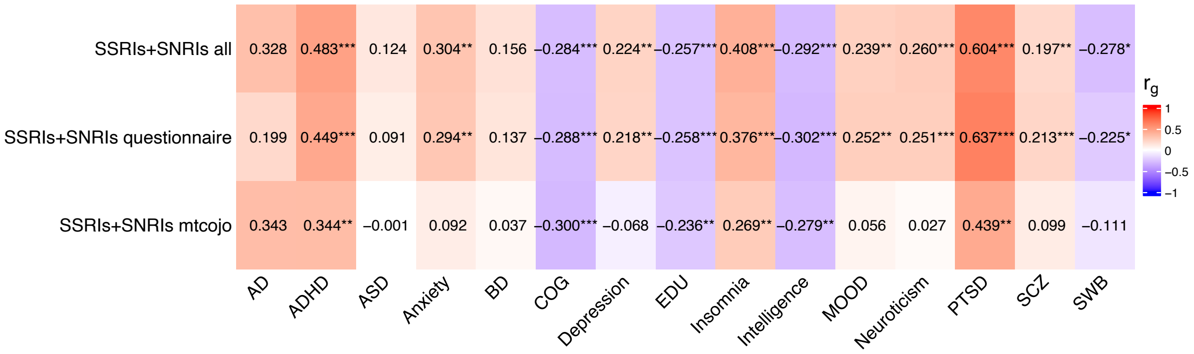


**Figure S8**: Genetic correlation between non-response SSRIs or SNRIs (from various meta-analyses) and Alzheimer´s disease (AD), attention deficiency hyperactivity disorder (ADHD), autism spectrum disorder (ASD), anxiety disorder, bipolar disorder (BD), cognitive performance (COG), educational attainment (EDU), insomnia, intelligence, depression phenotypes, mood instability (MOOD), neuroticism, posttraumatic stress disorder (PTSD), schizophrenia (SCZ), subjective well-being (SWB). *p<0.05, **p<0.01, ***p<0.001

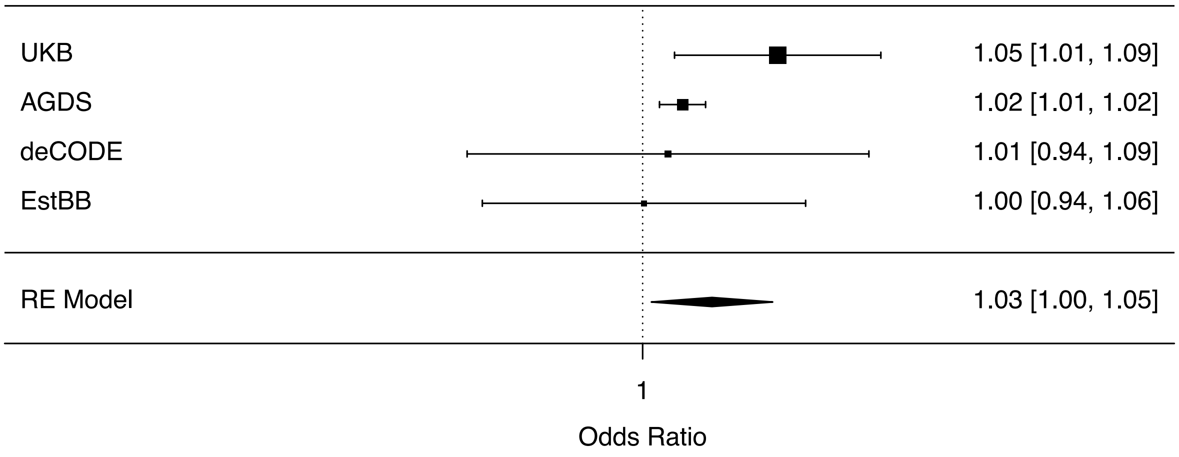


**Figure S9**: Forest plots showing the results from leave-one-out polygenic prediction of non-response to SSRIs in four independent cohorts, as well as meta-analyzed across these cohorts, weighted based on effective sample size. Effects are reported as odds ratios (95% confidence interval).

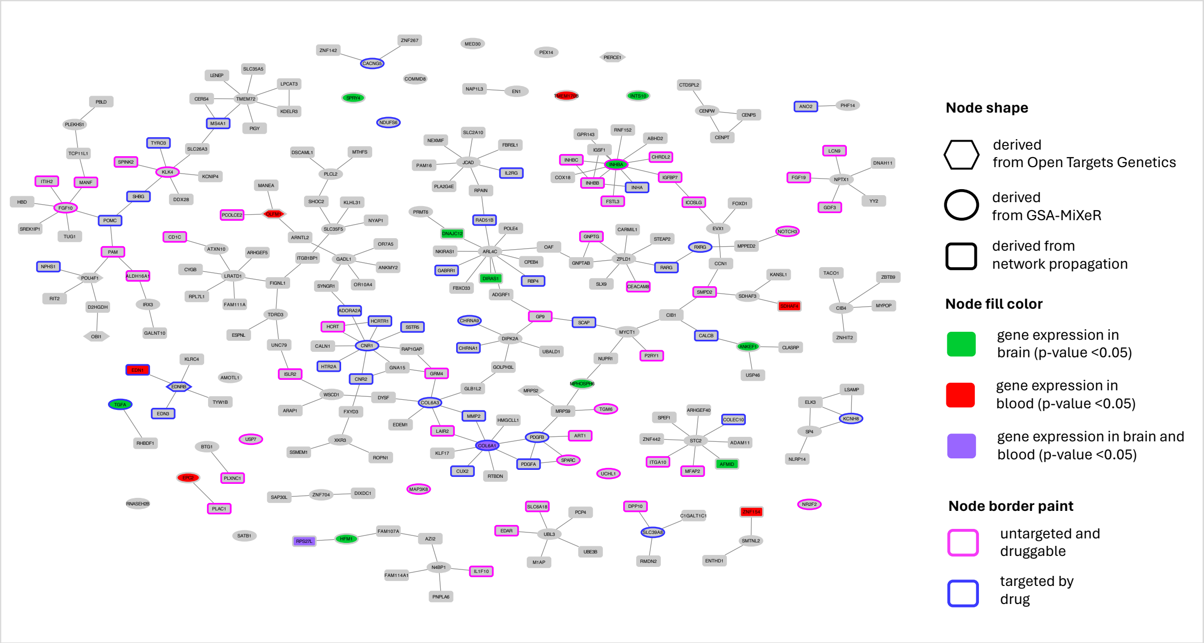


**Figure S10**: SSRI network. Nodes refer to genes, and edges refer to gene-gene interactions through identified protein-protein interactions between gene products (proteins).

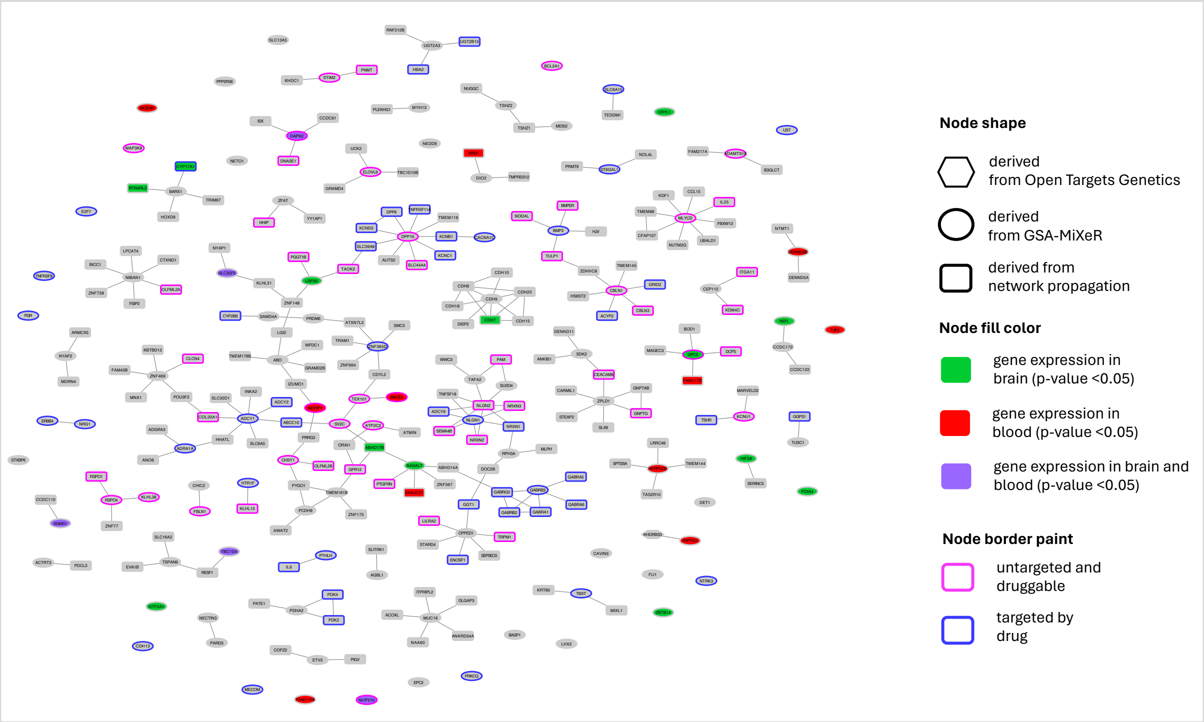


**Figure S11**: SNRI network. Nodes refer to genes, and edges refer to gene-gene interactions through identified protein-protein interactions between gene products (proteins).

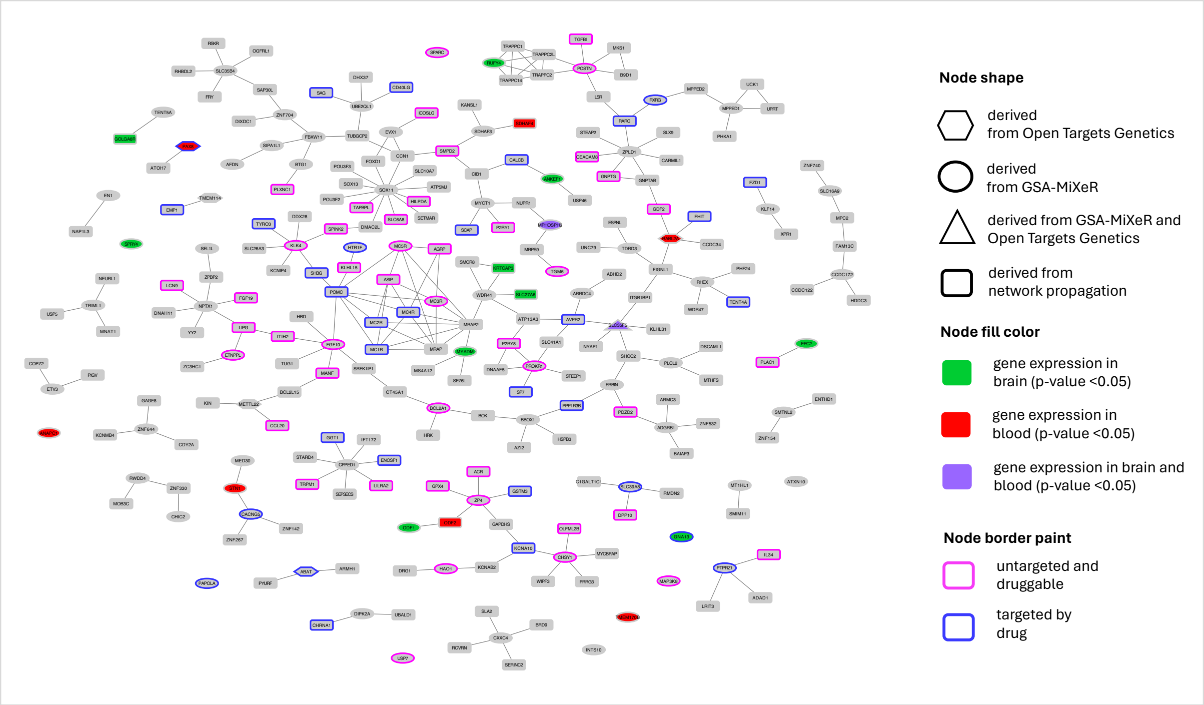


**Figure S12**: SSRI/SNRI network. Nodes refer to genes, and edges refer to gene-gene interactions through identified protein-protein interactions between gene products (proteins).

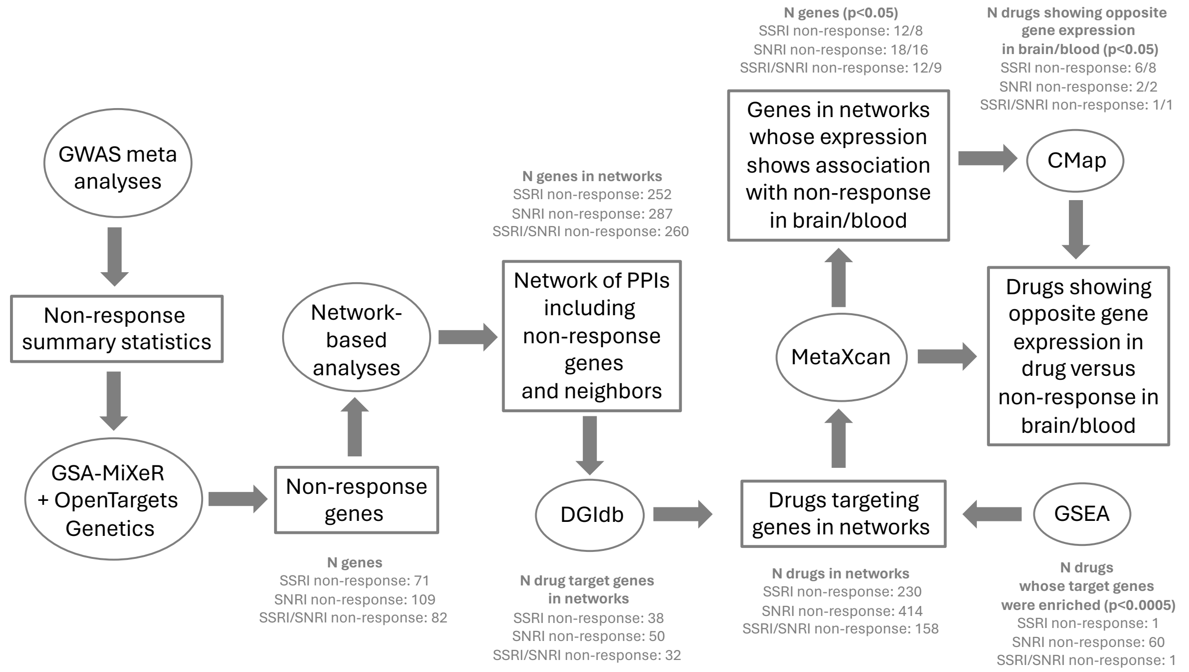


**Figure S13**: Flow chart depicting the steps taken in this study to identify drugs that could potentially address antidepressant non-response with corresponding numbers of genes and drugs.
